# Genetic Diversity of the Malaria Vaccine Candidate PfRIPR in a High Transmission Region of Senegal

**DOI:** 10.1101/2025.09.23.25335263

**Authors:** Megha Nair, Giselle Geering, Alyssa Agarwal, Rebecca Li, Yujie Qiao, Qin Xiao, Mariama N. Pouye, Laty G. Thiam, Aboubacar Ba, Kelly A. Hagadorn, Awa Cisse, Noemi Guerra, Yome Tawaldemedhen, Khadidiatou Mangou, Adam J. Moore, Fatoumata Diallo, Seynabou D. Sene, Bacary D. Sadio, Elizabeth Zhang, Lawrence Shapiro, Saurabh D. Patel, Alassane Mbengue, Ines Vigan-Womas, Zizhang Sheng, Amy K. Bei

## Abstract

RH5-Interacting Protein (RIPR) is essential for the invasion of *Plasmodium* into host red blood cells and is currently being studied as a novel malaria vaccine candidate in Phase 1a clinical trials. To study the genetic diversity of RIPR, deep amplicon sequencing was used to identify RIPR mutations in *Plasmodium falciparum* clinical isolates (n=89) collected in Kédougou, a high malaria transmission region of Senegal. We identified nonsynonymous single nucleotide polymorphisms (SNPs) in 64/89 (71.9%) of the samples. In total, 26 non-synonymous SNPs were identified, of which 15 were novel. 16/26 SNPs were able to be threaded onto existing RIPR crystal structures to predict the effects of SNPs on RIPR stability. 7/16 mutations were predicted to destabilize RIPR while 2/16 increased the stability of RIPR. Additionally, we identified 3 SNPs (Q737K, T738K, V840L) in the EGF5-8 domains of RIPR where neutralizing antibodies are known to bind.

## Introduction

Malaria is a mosquito-borne disease caused by parasites of the genus *Plasmodium*. While improved vector control measures (insecticides, bed nets) and chemoprophylaxis have led to progress in recent decades, malaria cases and deaths have spiked again in recent years. In 2023, there were 263 million cases of malaria and 597,000 malaria deaths with 95% of deaths occurring in Africa, disproportionately impacting children under five^1^.

To date, there are two WHO-approved malaria vaccines: RTS,S/ AS01 and R21/Matrix-M. These vaccines target the circumsporozoite protein (CSP) expressed during the sporozoite stage of the parasite life-cycle. However, these vaccines have limited efficacy with low antibody titers and strain-specificity. This poses an issue to vaccine success because the escape of a single sporozoite can result in the release of merozoites into the bloodstream which will not be targeted by vaccine elicited immunity. These merozoites undergo multiple rounds of asexual replication in erythrocytes, causing the clinical symptoms of malaria. Thus, recent efforts have focused on the development of a blood-stage vaccine to complement approved pre-erythrocytic vaccines^2^.

The five-membered PCRCR complex (consisting of PTRAMP, CSS, RIPR, CyRPA, and RH5) has emerged as leading target blood-stage antigens for a next-generation vaccine^3,4^. The PCRCR complex binds to basigin and is essential for *Plasmodium falciparum* invasion of red blood cells. A RH5 vaccine (RH5.1/Matrix M) has entered a phase 2b clinical trial in children in Burkina Faso^5^. The vaccine is able to elicit strong neutralizing antibodies in study participants. A combined RIPR-CyRPA vaccine (R78C) has reached Phase 1a clinical trials and is able to induce antibodies that are highly neutralizing against lab strains of *Plasmodium falciparum*. However, additional information on the genetic polymorphisms of RIPR is required to understand the potential for immune evasion and selection to optimize this antigen for a vaccine. We have previously examined the genetic diversity of PfRH5^6,7^ and PfCyRPA^8^ and the genotype/phenotype relationship between RH5 mutations and susceptibility to RH5 vaccine-elicited antibodies^9^.

Past efforts to develop a blood-stage vaccine have proven difficult given the redundancy of parasite invasion ligands (eg: EBAs) and the high rates of polymorphisms leading to strain-specific efficacy (e.g.: MSP-1^10^ and AMA-1^11–13^). To address this challenge and to generate a better understanding of the impact of PfRIPR genotype on potential functional outcomes for either merozoite invasion or immune evasion, we utilize deep amplicon sequencing to examine the genetic diversity of PfRIPR in 89 patient blood samples collected from Kédougou, a high malaria transmission region of Senegal. We also utilize structure modeling to determine the impact of the identified mutations on RIPR stability and the binding of neutralizing antibodies.

## Results

### Study Participants

This study was conducted in Kédougou, a region in the southeastern part of Senegal which has high malaria transmission during the rainy season (May-November). Blood samples from 89 participants who had confirmed *Plasmodium falciparum* infections were included from 2019 (n=15), 2022 (n=31), and 2023 (n=43). Study participants were recruited with informed consent from healthcare sites in six locations within Kédougou: Bandafassi, Bantako, Camp Militaire, Dalaba, Mako, and Ndiormi. **Table 1** lists the demographic and parasitological characteristics of the participants included. In our study population, we observed an overall complexity of infection (COI) of 2.71 and a sex ratio of 1.1. COI indicates the number of genetically distinct parasite strains present in each sample as determined by merozoite surface protein (MSP) genotyping. Participants enrolled in this study were aged 2 to 67 years (Median 17; SD = 14.66), and there were 45 males and 41 females. While there was not a striking difference in age distribution across sites, there was a lower proportion of children (≤10 years) at each site.

**Table 1.** Socio demographic and parasitological characteristics of study participants. Socio-demographic and parasitological characteristics of study participants. P-values were calculated using one-way ANOVA analysis on the GraphPad Prism version 10.5.0.

| Sites | BF | BANT | CM | DAL | MAK | NDI | Total | p-value |
| --- | --- | --- | --- | --- | --- | --- | --- | --- |
| Patients, No | 8 | 4 | 7 | 47 | 2 | 21 | 89 |  |
| Sex ratio (M/F) | 0.75 | 3 | 2.5 | 1 | 2 | 0.82 | 1.1 | 0.67* |
| Age (median, years) | 19 | 14 | 15 | 19 | 12 | 14 | 17 |  |
| [min-max] | [12–26] | [2–33] | [2–35] | [4–67] | [7–17] | [4–50] | [2–67] |  |
| $\leq 10$ years (%) | 0 | 25 | 14.29 | 8.69 | 50 | 33.33 | 16.09 | |
| $> 10$ years (%) | 100 | 75 | 85.71 | 91.31 | 50 | 66.67 | 83.91 | |
| COI (mean) | 2.25 | 4.33 | 6 | 2.46 | 6.5 | 1.8 | 2.71 | 0.0016** |
| [min-max] | [1–5] | [3–5] | [2–15] | [1–5] | [4–9] | [1–3] | [1–15] |  |
| $\leq 10$ years (mean) | N/A | N/A | 3 | 2.75 | 4 | 1.6 | 2.36 | |
| $> 10$ years (mean) | 2.43 | 4.33 | 6.75 | 2.44 | 9 | 1.9 | 2.80 | |

\* Statistical differences were calculated using the Chi-square test
\*\* Statistical differences were calculated using the Kruskal Wallis test
BF – Bandafassi, BANT – Bantako, CM – Camp Militaire, DAL – Dalaba, MAK – Mako, NDI – Ndiormi

### Identification of Non-Synonymous Single Nucleotide Polymorphisms (SNPs) in PfRIPR

A total of 89 *P. falciparum* positive blood samples were analyzed by deep amplicon sequencing to identify nonsynonymous single nucleotide polymorphisms (SNPs) in PfRIPR with a minimum variant read frequency of 2%. The PfRIPR sequence of 3D7 (*PF3D7_*0323400) was used as the reference allele. Of our samples, 25/89 (28.1%) had the 3D7 reference allele and nonsynonymous SNPs were identified in the remaining 64/89 samples (71.9%). We identified 26 non-synonymous SNPs in these samples of which 11 had been previously described in other datasets (A19E, V190A, N215K, Y259H, M255I, T327A, I373L, S552N, Q737K, Y985N, I1039M)^14–17^. Four of the previously identified SNPs were also the most prevalent SNPs in our dataset: Y259H (37/89, 41.6%), V190A (21/89; 23.6%), M255I (4/89; 4.5%), and Y985N (4/89; 4.5%) **(Figure 1A)**. We identified 15 novel SNPs in our study: D106N, V163F, H191N, A197S, D262H, E321*, C348F, V414L, D454G, K485N, D596Y, P716Q, T738K, V840L, S1010*. All SNPs were only identified in a single sample, with the exception of D454G which was identified in two samples. Of the samples with SNPs, 41/64 (64.1%) had one SNP, 16/64 (25%) had two SNPs, and 7/64 (10.9%) had three SNPs. The combinations of SNPs detected in samples are shown **(Figure 1A)**. The most common SNP combination was Y259H + V190A (5 samples), and Y259H + S552N (3 samples). Other combinations were only found in a single sample each.

**Figure 1:**
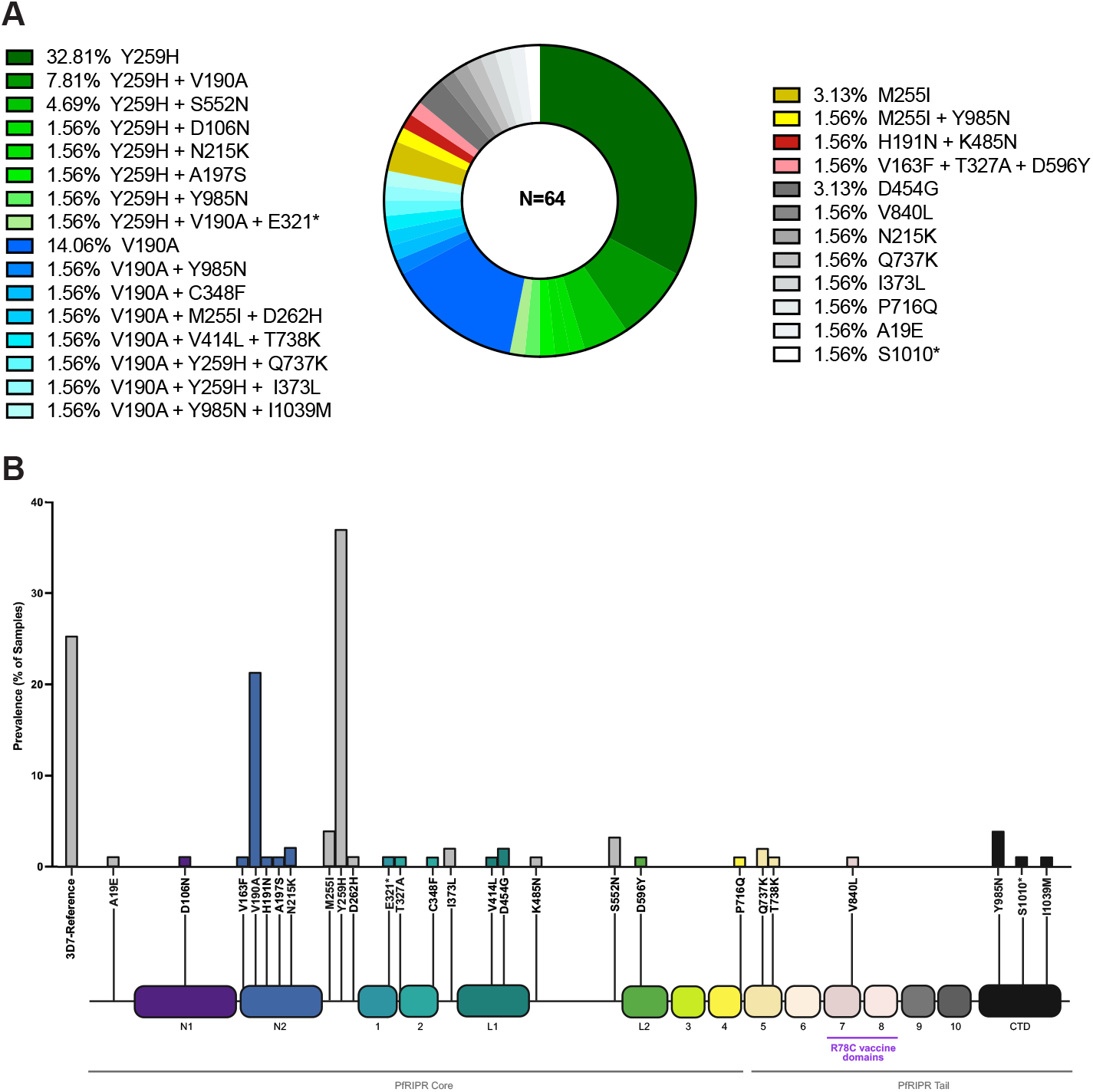
Identification of Non-Synonymous Single Nucleotide Polymorphisms (SNPs) in PfRIPR. **A)** All PfRIPR non-synonymous single nucleotide polymorphisms (SNPs) identified in the samples included in the study. Percentages refer to the percent of samples with a given SNP (or combination of SNPs) out of the total number of samples which have a SNP (n=64). **B)** Relative Position of Each Non-Synonymous SNP in the PfRIPR Gene. Nonsynonymous single nucleotide polymorphisms (SNPs) in PfRIPR for 89 samples. Reference allele refers to the PfRIPR sequence from the 3D7 reference genome (*PF3D7_*0323400). Below the bar chart is a schematic representation of the PfRIPR gene indicating the positions of all non-synonymous SNPs identified in this study. Boxes denote the domains of PfRIPR, including ten epidermal growth factor-like (EGF) domains. The EGF5-EGF8 domains include growth-inhibitory antibody epitopes. The EGF7 and 8 domains included in the R78C vaccine are labelled^18^.

Two of the identified SNPs resulted in truncation of the RIPR protein at amino acid positions 321 and 1,010. Each SNP was only found in a single sample with E321* detected at a variant read frequency of 14.3% and S1010* at a frequency of 2.5%. As PfRIPR is an essential protein and the interaction of the PfRIPR tail domain with CSS and PTRAMP has been shown to be necessary for invasion, these SNPs likely do not represent functional proteins within the parasite.

The relative position of each non-synonymous SNP in the PfRIPR gene is shown in a simplified schematic **(Figure 1B)**. Only 3 SNPs (Q737K, T738K, V840L) were identified in the epidermal growth factor-like (EGF) domains 5-8 (AA 719-897). Antibodies binding to the EGF5-EGF8 domains have previously been shown to have neutralizing activity in the growth inhibition assay. Thus, it is possible that these three SNPs may alter antibody binding and lead to immune evasion^18^.

### Variant Read Frequency of Non-Synonymous PfRIPR SNPs

We next examined the variant read frequency for each of the SNPs we identified **(Supplemental Figure 1)**. For a given sample, variant read frequency represents the percent of reads at a position which have a SNP. In high transmission settings such as the study site of Kédougou, mixed-genotype malaria infections are common. Out of 89 clinical samples reported here, only 17 samples were monogenomic infections (COI = 1). In our dataset, the majority of samples were polygenomic infections, and the overall COI values ranged from 1 to 15, with a mean of 2.71 genotypes per sample.

In our data, there was a large range of variant read frequencies within samples (Supplemental Figure 1). All 11 of the previously identified SNPs had high variant read frequencies. The two most common SNPs (Y259H and V190A) both had high variant read frequencies (mean variant read frequency for Y259H=66.5% and V190A=65.9%). However, each SNP also had very large variation in the variant read frequency across samples. Of the 15 novel SNPs identified in this study, 9/15 SNPs (H191N, A197S, C348F, K485N, D596Y, P716Q, T738K, V840L, S1010*) were present at low variant read frequency (2-5%), 3/15 were present at intermediate variant read frequency (5-25%) (D106N, E321*, and V414L), and 3/15 SNPs were present at high variant read frequency (>25%) (V163F, D262H, D454G). Of the three SNPs (Q737K, T738K, V840L) identified in the PfRIPR neutralizable domains (EGF domains 5-8), Q737K had a high variant read frequency (99.3% and 56.7%) in both samples **(Supplemental Figure 1)**. However, both T738K and V840L had low variant read frequencies (3.6% and 3.7% respectively).

Out of the 17 samples with monogenomic infections, 12 carried at least one mutation on the PfRIPR gene. Eight samples carried Y259H, with variant read frequencies ranging from intermediate to high (9.2%, 24.1%, 99.9%, 100%, 100%, 100%, 100%, and 100%). V190A was present in 4 samples at intermediate to high frequencies (44.2%, 81.1%, 100%, 100%), and 2 carried S552N at high variant read frequencies (54.6% and 100%) **(Supplementary Table 4)**.

### Structural Modeling of SNPs

We next determined the effects of the identified SNPs on the stability of RIPR and the RIPR/CyRPA interaction **(Table 2)**. A cryo-EM structure of PfRIPR in complex with PfRH5 and PfCyRPA has been published^19^. However, the cryo-EM map this model is based on did not have density for PfRIPR beyond residue 716. Consequently, there currently only exists a low-confidence AlphaFold2 prediction of the PfRIPR tail (residues 717-1086)^20^. This elongated tail is the site of R78C vaccine domains EGF7 and EGF8 and is of particular interest to vaccine immunogen design because it is the target of growth neutralizing antibodies. However, due to the low resolution of the tail domain, we were unable to thread SNPs located on EGFs 5-8 onto the PfRIPR structure^19^. For each of the 16 SNPs we were able to thread onto the structure of PfRIPR, we looked their effects on the stability of RIPR alone and the stability of the RIPR/CyRPA interaction. The result showed that 7/16 were destabilizing mutations and 2/16 were stabilizing mutations. The top destabilizing mutations tend to show low circulating frequencies and low co-occurrence.

**Table 2:** Structural Effects of PfRIPR SNPs. Out of 24 SNPs, 16 were threaded onto existing PfRIPR crystal structures. The effect of SNPs on RIPR stability and RIPR-CyRPA binding energy were predicted by FoldX 4.0. A mutation was considered stabilizing if the change in stability was less than -0.500 kcal/mol (highlighted in green). A destabilizing mutation refers to a change in stability greater than +0.500 kcal/mol (highlighted in red).

| SNP | $\Delta\Delta G$ RIPR/CyRPA Stability (kcal/mol) | $\Delta\Delta G$ RIPR Stability (kcal/mol) |
| --- | --- | --- |
| N56D | 0.000 | 0.495 |
| N66Y | -1.287 | 0.471 |
| D106N | 0.000 | -0.034 |
| G119A | 0.000 | 3.922 |
| V163F | -0.010 | 0.109 |
| I373L | 0.000 | -0.381 |
| V190A | 1.206 | 2.628 |
| H191N | 0.731 | -0.561 |
| Y184S | 0.001 | 1.315 |
| A197S | 0.126 | 0.549 |
| L199S | 0.020 | 3.604 |
| N215K | 0.000 | 0.423 |
| Y226H | 0.007 | 3.267 |
| F236V | -0.047 | 2.807 |
| M255I | 0.000 | 0.022 |
| Y259N | 0.000 | 0.499 |
| Y259H | 0.000 | 0.954 |
| D262H | 0.003 | 1.657 |
| T327A | 0.000 | -0.745 |
| C348F | 0.000 | 13.282 |
| G341R | 0.000 | 5.693 |
| V414L | 0.000 | 0.617 |
| V429L | 0.000 | 0.363 |
| M436V | 0.000 | 1.903 |
| D438N | 0.000 | -0.286 |
| D438G | 0.000 | -0.537 |
| T441N | 0.000 | 0.071 |
| A444T | 0.000 | 2.370 |
| D454G | 0.000 | -0.499 |
| D596A | 0.000 | -0.014 |
| D596Y | 0.000 | 0.154 |
| L673V | 0.000 | 1.137 |
| P716Q | 0.000 | 0.884 |

Y259H was the most prevalent SNP in our dataset (37/89, 41.6%), and in our structural analysis it exhibited a destabilizing effect on PfRIPR with a ΔΔG of +0.954 kcal/mol but had no effect on the stability of the PfRIPR/PfCyRPA interaction **(Table 2)**. Y259 forms a hydrogen bond with Q181, which forms a hydrogen bond network with E59 and D61 **(Figure 2B)**. This hydrogen bond network plays an important role to stabilize the interaction between EGF1 and N2. Y259H disrupts the hydrogen bond between 259 and 181 and destabilizes the inter-domain interaction. In some cases, protein function may be enhanced by increased flexibility at the expense of stability^21^. Past three-dimensional variability analysis of cryo-EM data has revealed flexibility in PfRIPR, particularly in the region which links its multidomain core to the tail. The PfRIPR tail mediates the binding of PfRIPR to PfCSS-PfPTRAMP, which positions the PCRCR complex for binding to basigin. If Y259H causes destabilization of PfRIPR, the resulting increase in flexibility may offer some advantage when PfRIPR orients itself to bind to PfCSS-PfPTRAMP and positions PfRH5 towards the erythrocyte for binding to basigin^19^.

**Figure 2:**
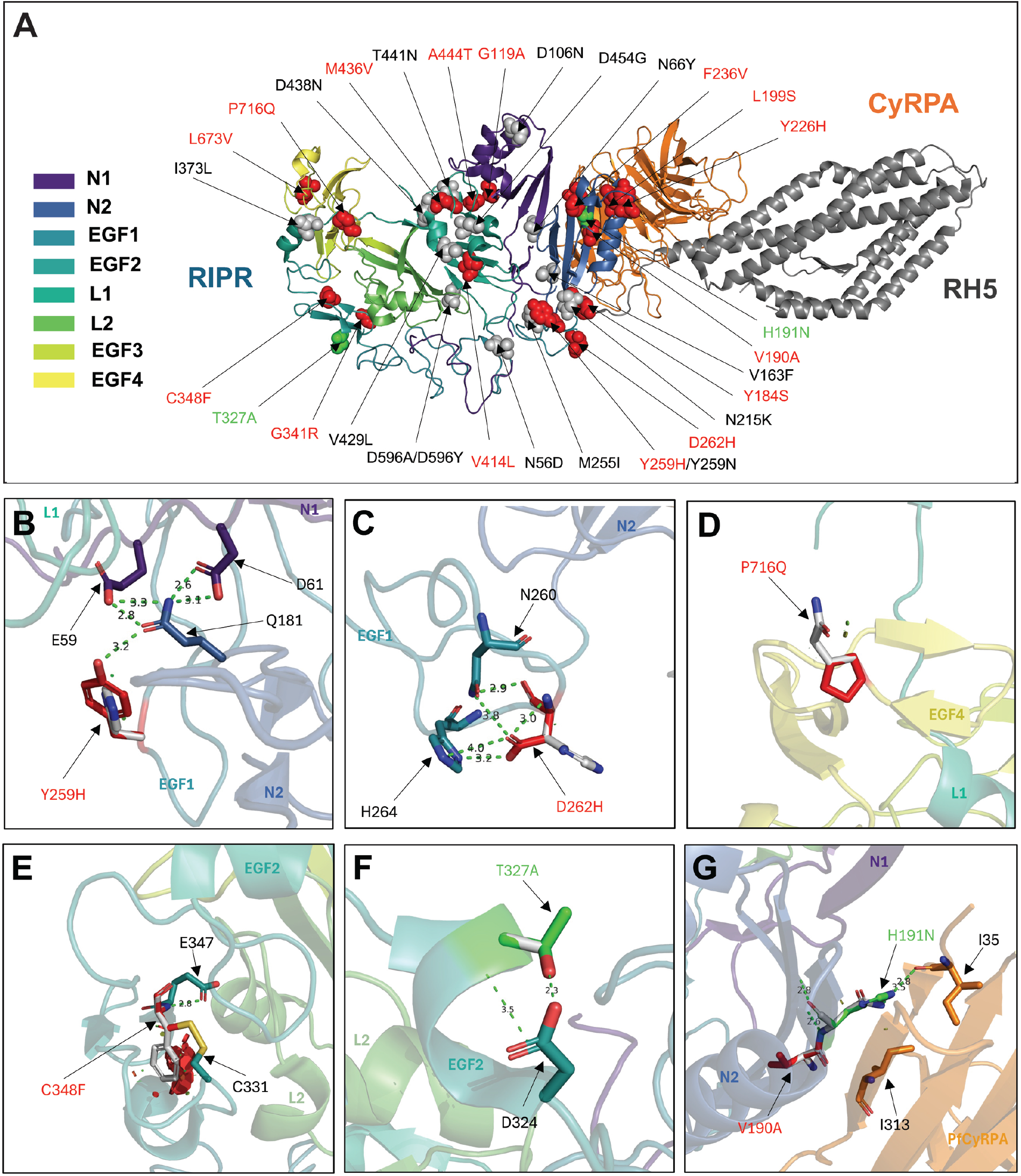
Structure of RH5-CyRPA-RIPR complex with threaded identified SNPs. **A)** The domains of PfRIPR are color-coded. Destabilizing SNPs are highlighted in red and stabilizing SNPs are highlighted in green. SNPs identified in this study are A19E, D106N, V163F, V190A, H191N, A197S, N215K, M255I, Y259H, D262H, T327A, C348F, I373L, V414L, D454G, K485N, S552N, D596Y, P716Q. Other SNPs identified in MalariaGEN are: N66Y, Y184S, L199S, G341R, D438N, H511R, H524L, L673V. SNPs identified in Ndwiga *et al* ^14^ include G119A, Y259N, V429L, M436V, D438G. SNPs identified in Ntege *et al*^15^ are N56D, A444T, D596A. SNPs identified in Waweru *et al*^16^ are Y226H, F236V, T441N. Close-up of side chain interactions with **B)** Y259H **C)** D262H **D)** P716Q **E)** C348F **F)** T327A **G)** H191N and V190A.

Along with Y259H, the SNPs D262H and P716Q **(Figure 2C and D)** also had destabilizing effects on PfRIPR, with a ΔΔG of +1.657 kcal/mol and +0.884 kcal/mol, respectively. D262H disrupts hydrogen bonds with H264 and N260, which increases the flexibility of the EGF1 loop **(Figure 2C)**.The SNP P716Q is intriguing because it is located at the very end of EGF4, the domain from which the PfRIPR tail extends from the RIPR core. Interestingly, residue 716 is the final residue in the PfRIPR core for which there is model density^19^. Thus, the SNP P716Q is positioned exactly before the start of domains EGF5-8, which encompass the PfRIPR tail and one of the most immunogenic sites of the RIPR protein. Since proline usually rigidify local structures, P716Q will enhance the flexibility between the PfRIPR core and EGF5-8. Thus, P716Q may have important consequences for parasite invasion, as the PfRIPR tail alone has been shown to be sufficient for the binding of PfRIPR to the PfCSS–PfPTRAMP complex, an interaction which is crucial for the subsequent binding of PfRH5 and PfCyRPA to basigin on the erythrocyte surface^19^.

Mutation C348F had by far the largest destabilizing effect on RIPR, with a ΔΔG of +13.282 kcal/mol **(Table 2)**. Position 348 is buried at the interior of domain EGF2. C348F destabilizes EGF2 through two mechanisms: substantial steric clashes introduced by the phenylalanine side chain and disruption of the disulfide bond between C348 and C331 **(Figure 2E)**. On the other hand, T327A was one of the two SNPs which had a stabilizing effect on PfRIPR with a ΔΔG of - 0.745 kcal/mol **(Table 2)**. T327, located at the middle of EGF2 helix, forms hydrogen bonds with D324 at the beginning of the helix **(Figure 2F)**. T327A mutation disrupted the hydrogen bonds and may enhance the flexibility of the helix. This SNP is also located the aforementioned site of RIPR cross-links, but contrary to C348F, the mutation at residue 327 could offer additional stability to RIPR intra-protein associations.

The V190A and H191N mutations **(Figure 2G and H)** were the only two threaded SNPs to cause destabilization in the PfRIPR/PfCyRPA interaction, with a ΔΔG of +1.206 kcal/mol and +0.731 kcal/mol, respectively **(Table 2)**. Through domain N2 of its structured core, PfRIPR binds to blade 5 of PfCyRPA. A destabilization in this association could have important implications for the protein interaction network of the RCR complex. The interface of the PfRIPR/PfCyRPA complex is formed mostly through hydrophilic side chain interactions^19^. . H191 forms a hydrogen bond with I35 of PfCyRPA. H191N may disrupt the hydrogen bond and form steric clashes with I313 of PfCyRPA. The V190A mutation is also an amino acid change from the hydrophobic valine to the hydrophobic alanine. As valine has a larger side chain than alanine, the V190A polymorphism would result in less hydrophobic interactions within the N2 domain. In our dataset, V190A has a much higher prevalence (23.6%) than H191N (1.56%) **(Figure 2)**. The low prevalence of H191N, (identified in one patient sample) may suggest that it is not a favorable SNP for parasite invasion or survival. On the other hand, V190A is a previously identified SNP that occurs at a high population prevalence in Africa and globally^17^. V190A is also one of the most prevalent SNPs in our dataset (23.6%) and makes up one part of the most common SNP combination (Y259H + V190A). Out of the 16 SNP combinations we identified, half of them (8/16) included V190A. The destabilizing effect of V190A could perhaps offer some advantage to *P. falciparum* during invasion, as protein instability has been linked to increased flexibility^22^, and flexibility has been observed along the PCRCR complex during its presentation to basigin^19^.

## Discussion

Genetic variation in vaccine targets can be a significant hindrance to malaria vaccine success, as polymorphisms can confer evolutionary advantages to parasites such as drug resistance^23^ or the facilitation of erythrocyte invasion^24^. As recent data revealed that a combination vaccine involving the C-terminal EGF domains of RIPR and CyRPA (R78C) can produce antibodies with higher GIA activity than RH5 alone^18^, understanding how genetic variation impacts *Plasmodium falciparum* invasion will be crucial to malaria elimination efforts. This study investigated the genetic diversity of the malaria vaccine candidate PfRIPR in Kédougou, a high transmission region of Senegal. The ability of anti-PfRIPR antibodies to block *P. falciparum* invasion and growth has indicated the promise of this essential antigen as a key component of future antimalarial vaccines^25^. While PfRIPR is highly conserved, the possibility of mutation remains. Therefore, we set out to identify immunologically relevant polymorphisms^20^ in PfRIPR to inform vaccine design and the interpretation of vaccine trial results.

To reveal rare PfRIPR variants that could impact vaccine efficacy, we used a deep targeted amplicon sequencing approach using Illumina short-read sequencing, ideal for identifying rare SNPs in the context of highly polygenomic infections. We have previously reported^20^ significant polyclonality in the high-transmission region of Kédougou. In our current study, the mean COI was 2.7. Thus, in order to generate data with high accuracy and depth, we employed an NGS platform. Overall, 64/89 (71.9%) of the isolates contained at least one SNP in the PfRIPR gene relative to the 3D7 reference genome. A similar trend was observed for PfRH5 in previous genetic analyses^6^. However, the opposite was observed for PfCyRPA, for which the 3D7 reference allele represented the dominant allele in the sampled population^8^.

We identified 26 non-synonymous SNPs in these samples of which 11 had been previously described in other datasets (A19E, V190A, N215K, Y259H, M255I, T327A, I373L, S552N, Q737K, Y985N, I1039M)^14–17^. Four of the previously identified SNPs were also the most prevalent SNPs in our dataset: Y259H (37/89, 41.6%), V190A (21/89; 23.6%), M255I (4/89; 4.5%), and Y985N (4/89; 4.5%). Additionally, we identified two truncations (E321* and S1010*) present in one sample each. As previously described, RIPR is an essential merozoite surface protein and conditional knock-outs have proved to be lethal to the parasite^3^, and further, the interaction of its tail region (EGF5-10) with PTRAMP and CSS is essential for its function. A mutation such as E321* that results in the truncation of PfRIPR would cause the protein to lack these important domains and would therefore be deleterious. In the *P. falciparum* life cycle, purifying selection has been shown to work efficiently^26^. This means that most segregating mutations present in the parasite genome are either neutral or very new and have not been weeded out by purifying selection, as is likely in the case of E321*. The other mutation which results in a truncation event occurs at amino acid 1,010, which occurs at the extreme c-terminus after EGF10, but is still within the tail region of PfRIPR. One possibility for our identification of two truncations of PfRIPR is that our very sensitive VAF threshold for detecting SNPs and high density of coverage may reveal rare SNPs in the process of being cleared by purifying selection. If we increase our SNP threshold to 5%, most of these SNPs would not be called. Interestingly, past genomic analysis of PfRH5 and PfCyRPA did not identify any polymorphisms that cause truncation events in these two components of the RCR complex. However, these truncations in PfRIPR warrant further functional investigation.

A past genetic study in Western Kenya also identified the SNPs Y259H, V190A, and M255I as high frequency in the Lake Victoria parasite population^16^. In another study using patient samples from Kilifi, Kenya, Ndwiga and colleagues also reported Y259H and V190A as high frequency SNPs within the RIPR gene, as well as Y985N and I1039M^14^. To describe genetic polymorphisms, they used whole genome sequencing data (WGS) and capillary sequencing data (CS).

Data generated from the MalariaGEN Pf8 data release revealed that three of the top four most prevalent SNPs in our dataset Y259H (41.6%), V190A (23.6%), and Y985N (4.5%) were also included among the top four most prevalent SNPs globally^17^. Notably, Y259H, V190A, and Y985N displayed a global prevalence, having been identified in samples across Africa, Asia, and Oceania, with Y259H and V190A showing a particularly high number of samples in West Africa. Of the SNPs located in EGF domains 5-8, only Q737K was reported in the MalariaGEN database as being present in samples from West Africa (n=22) and East Africa (n=6). Further population-specific studies across additional endemic regions could generate a more holistic pool of knowledge for future immunogen design.

The RIPR protein has 10 epidermal growth factor-like (EGF) domains. Previous studies have strongly suggested epitopes located in the “tail” region domains EGF5-EGF8 (AA 719-897) are likely the sole target of growth inhibitory antibodies^18^. Only 3 SNPs (Q737K, T738K, V840L) identified in our study are located in the EGF domains 5-8. Of particular interest is V840L, which is located in one of the R78C vaccine domains, EGF 7. As previously described, the lack of crystal structure data for the tail of PfRIPR prevented the structural threading of these SNPs. Once available, structural data for PfRIPR will be crucial to understanding the functional impact of SNPs on immune evasion. In addition, defining the antigenic landscape of PfRIPR will help us understand how identified polymorphisms impact the binding of neutralizing human monoclonal antibodies from R78C-vaccinated individuals. Currently, no work has been published, to our knowledge, that identifies the specific epitopes targeted by leading anti-PfRIPR antibodies, beyond EGF domains and regions within PfRIPR. Future studies aiming to define the epitope landscape of PfRIPR and map these epitopes onto its structure could further elucidate the effect of identified SNPs on immune evasion. For example, epitope binning data has been generated for both PfRH5 and PfCyRPA^27^, and for PfRH5 in particular, a single polymorphism (S197Y) was shown to impact the binding and function of the mAb R5.017 to PfRH5^18,28^. Defining the antigenic landscape of PfRIPR in a similar manner can strengthen our understanding of how polymorphisms identified in PfRIPR impact mAb binding and function, thus refining vaccine immunogen design.

To further understand the functional impacts of identified SNPs, we threaded 16 SNPs onto the core of PfRIPR. Seven of the 16 SNPs were destabilizing mutations and 2/16 were stabilizing mutations. A recurring consideration during the analysis of these SNPs was that of RIPR tail flexibility and function. Increased protein flexibility has been shown to enhance protein function at the expense of stability, meaning that a SNP which destabilizes the RIPR protein could potentially result in increased flexibility to bind other proteins. For example, P716Q was shown to have a destabilizing effect on PfRIPR, and its position at the end of EGF4 and the beginning of the PfRIPR tail suggests that this SNP has an impact on stability of the PfRIPR tail and thus influences how the tail can bind to the PfCSS–PfPTRAMP complex before ultimately orienting RH5 and CyRPA to bind basigin. Another destabilizing SNP, C348F, had by far the greatest destabilizing effect on RIPR, with a ΔΔG of 13.282 kcal/mol, a fivefold higher ΔΔG than the next largest destabilizing SNP, V190A, at 2.628 kcal/mol. The significant destabilizing effect of C348F on RIPR is likely attributed to its position in EGF2. The mutation of residue 348 from cysteine to the bulkier phenylalanine could lead to steric clashes and disrupt the disulfide bond between C348 and C331. The low frequency of this mutation implies that C348F may be a deleterious mutation that has not been weeded out by purifying selection.

The V190A mutation was one of the two threaded SNPs to cause destabilization of the PfRIPR/PfCyRPA complex. Not only is V190A a previously identified SNP, but it is also one of the most prevalent SNPs in our dataset (23.6%) and makes up one part of the most common SNP combination (Y259H + V190A). V190A is located on the interface between PfRIPR and PfCyRPA. Past data has shown that blocking the formation of the RCR complex, such as the PfRIPR/PfCyRPA interface, is not an inhibitory mechanism^27^. Thus, the PfRIPR/PfCyRPA interface is not a priority target for vaccine design. While this interface is not an attractive inhibitory domain, the PfRIPR/PfCyRPA complex — particularly the R78C formulation — has shown to improve overall GIA when combined with RH5.1^27^. Therefore, understanding the destabilizing effect of V190A on the PfRIPR/PfCyRPA complex could be important to future studies of anti-CyRPA and anti-RIPR antibody combinations which have shown promising intra-antigen synergistic GIA data. An additional consideration is that of compensatory mutations that could potentially exist within the PCRCR complex. A destabilizing mutation in one protein may be balanced out by compensatory mutations in other members of the complex. Combining genomic data across the PCRCR complex, including the data generated in this study, can help us understand these potential compensatory effects.

While *P. falciparum* is the most widespread, five other species within the *Plasmodium* genus cause human malaria. A previous study found that RIPR-induced antibodies were cross-reactive when tested against sera from individuals infected with *P. falciparum, P. knowlesi*, or *P. vivax*^29^ . The RIPR orthologs found in these species are also essential for erythrocyte binding and associate with CSS and PTRAMP orthologs. Further studies that analyze genetic diversity of RIPR in other human *Plasmodium* species will further inform the design of a vaccine that is effective across species.

In this study, we employed deep targeted amplicon Next Generation Sequencing for short read Illumina sequencing. While highly sensitive, short read NGS does present some limitations. Of particular concern for this study is that the short reads generated can make it difficult to assemble full PfRIPR haplotypes within our polygenomic samples. Without knowledge of the sequences flanking each 150 bp fragment, we are unable to determine which strain of *P. falciparum* is associated with a given SNP. In future analysis, we will complement our short-read sequencing with long-read sequencing, such as Oxford Nanopore, which can generate complex, long reads of DNA which can distinguish full-length PfRIPR haplotypes in polyclonal infections, albeit with decreased sensitivity to uncover rare variants.

Another limitation of our study is the relatively small sample size of our data. The 89 patient samples we included were collected over the span of three years (2019, 2022, 2023) with an uneven distribution of patient samples across years. In addition, expanding the geographic regions in which the genetic diversity of PfRIPR is studied will create a more holistic pool of knowledge for future immunogen design. This means gathering data from other highly endemic regions across Africa, for example.

While our model provides predictive data on the structural effects of identified polymorphisms, validation will require biochemical studies of antibody-protein binding and phenotypic assays. Future work will use *in vitro* phenotypic assays to investigate genotype-phenotype associations and the impact of SNPs on parasite invasion and susceptibility of the parasite to vaccine-induced antibodies. For example, functional growth inhibition assays (GIAs) will be used to measure the ability of neutralizing antibodies to inhibit the growth of CRISPR/Cas9-generated transgenic parasites expressing SNPs of interest.

RIPR is an essential merozoite protein and vaccine target that has been shown to be highly immunogenic in animal and human immunizations^27^. From our study utilizing *P. falciparum* samples from the malaria-endemic region of Kédougou, Senegal, we conclude that RIPR is a highly conserved antigen with most of the identified mutations having destabilizing effects in RIPR alone, but minimal impact on the RIPR-CyRPA interaction, and most are localized outside of known neutralizing antibody-binding regions. Additional phenotypic studies must be conducted to determine the effect of polymorphisms on natural and vaccine-induced antibodies. Developing improved malaria intervention strategies will rely on integrating epidemiologic, structural, and genetic data from malaria-endemic geographic regions.

## Data Availability

The sequencing data generated by this study have been deposited in the NCBI SRA under the following BioProject accession number: PRJNA1204383

https://www.ncbi.nlm.nih.gov/bioproject/PRJNA1204383

## Resource Availability

### Lead Contact

Requests for further information and resources should be directed to the lead contact, Amy Bei.

### Materials Availability

No new materials were generated by this study.

## Acknowledgements

This work was supported by the Fogarty International Center of the NIH (K01 TW010496), National Institute of Allergy and Infectious Diseases of the NIH (R01 AI168238), and G4 group funding (G45267, Malaria Experimental Genetic Approaches & Vaccines) from the Institut Pasteur de Paris and Agence Universitaire de la Francophonie (AUF) to A.K.B. This work was also supported by National Institute of Allergy and Infectious Diseases of the NIH (R61 AI176583) to Z.S. This work has been produced with the financial assistance of the European Union (Grant no. DCI-PANAF/2020/420-028), through the African Research Initiative for Scientific Excellence (ARISE), pilot program. ARISE is implemented by the African Academy of Sciences with support from the European Commission and the African Union Commission. The contents of this document are the sole responsibility of the authors and can under no circumstances be regarded as reflecting the position of the European Union, the African Academy of Sciences, and the African Union Commission. Additional funding for student exchange and research was provided by the Yale School of Public Health Wilbur Downs Fellowship (M.N., K.A.H., Y.T.), the Yale Collaborative Action Project Fellowship (K.A.H., Y.T., N.G., M.N., G.G.), the Yale Lindsay Fellowship for Research in Africa (K.A.H, Y.T.), the Ambrose Monell PhD Research Fellowship (K.A.H.), the Yale Tetelman Fellowship (A.C., R.L., N.G.), the Yale Richter Summer Fellowship (A.C., N.G.), the Yale Global Health Awards for Research (N.G.), and the Yale College Dean’s Office Perspectives on Biological Research Fellowship (to G.G.).

We are grateful for the support of the following colleagues from our field sites: Souleymane Ngom from Dalaba, Lt. Dr Charles Latyr Diagne and Lamine Kane from Camp Militaire, Moctar Mansaly, Adama Diallo, and Gerald Keita from Bandafassi, Safietou Sane and Astou Ndiaye from Mako, Adama Gueye from Bantako, and Dr. Ababacar Leye from Ndiormi. Additionally, we would like to thank all the healthcare workers at these sites for their partnership with Institut Pasteur de Dakar. We would also like to thank the people and communities of Kédougou for their invaluable contributions to this work. We are grateful for the support of the Yale Center for Genome Analysis (YCGA) for their support with sequencing. This publication uses MalariaGEN data as described in ‘Pf8: an open dataset of Plasmodium falciparum genome variation in 33,325 worldwide samples’ MalariaGEN et al, Wellcome Open Research 2025, 10:325 https://doi.org/10.12688/wellcomeopenres.24031.1.

## Author Contributions

A.K.B. conceived the experiments. A.K.B., Z.S., and L.S. supervised the research. R.L., L.G.T., M.N.P., A.B., Y.T. A.C., N.G., K.H., K.M., A.J.M., F.D., S.D.S, A.T., B.D.S., and A.M. collected the samples. M.N., G.G., R.L., E.Z. and A.A., conducted the experiments. Y.Q., Q.X., S.D.P. and Z.S., performed structural modelling. M.N., G.G., A.A., M.N.P., and A.K.B. analyzed the sequencing data. M.N. and G.G. wrote the manuscript. A.K.B. and Z.S. reviewed and edited the manuscript. All authors reviewed and approved the final manuscript.

## Methods

### Sample Collection

Samples were collected in June and July of 2019 2022, and 2023 in Kédougou, a highly endemic rural area in southeastern Senegal where malaria transmission occurs during the rainy season (May to November). The study protocol was approved by National Ethics Committee of Senegal (CNERS) (SEN19/36 and SEN23/09), the regulatory board of the Senegalese Ministry of Health and the Institutional Review Board of the Yale School of Public Health (2000025417). Participants were identified through passive case detection at the following clinical sites: Dalaba (2019, 2022, 2023), Mako (2019), Bantako (2019), Ndiormi (2023), Camp Militaire (2019), and Bandafassi (2022). Patients who presented to the clinic with febrile illness (fever in the past 24 hours and a temperature of >38 C) and/or a confirmed diagnosis of a *Plasmodium falciparum* mono-infection by both rapid diagnostic test (RDT) and microscopy were eligible for the study. Informed consent was obtained from study participants and/or their legal guardians. 5 mL of venous blood was drawn from patients who gave consent and samples were transported to the research station for processing. All samples were de-identified and sample and patient IDs were not known to anyone outside the research group.

### PfRIPR Sequencing

The QIAamp DNA Blood Mini Kit (Qiagen, 51106) was used to extract DNA from dried blood spots (DBS). For 2022 and 2023 samples, the full length PfRIPR sequence was PCR amplified. For 2019 samples, PfRIPR was PCR amplified in two parts. The primers and protocols are described in **Supplementary Tables 1-3**. The size and purity of amplicons were confirmed on an agarose gel before proceeding with the Next Generation Sequencing (NGS) protocol. All PCR amplicons were bead-purified (Omega Biotek, M1378-01), quantified using Qubit, and adjusted to 0.25 ng/µL. Library preparation was completed with Nextera XT kit with DNA/RNA unique dual indexes (UDI) (Illumina). Samples were then bead-purified again and assessed by qPCR-based quantification using the KAPA Library Quantification Kit (Roche, 07960140001). All samples were then normalized to a concentration of 4 nM and pooled into 8 sub-pools. The sub-pools were again bead-purified, quantified by KAPA qPCR, and normalized to 4 nM. The normalized sub-pools were then combined to form the final pool. The final pool was submitted for paired-end 150 bp sequencing on the Illumina NovaSeq 6000 platform at the Yale Center for Genome Analysis (YCGA). At least 1,000,000 reads per sample were obtained. Reads were de-multiplexed to obtain forward and reverse sequencing reads for each sample. For each sample, reads were uploaded to Geneious Prime, then paired and trimmed with the BBDuk plug-in. Trimmed reads were mapped to the RIPR sequence from the Pf3D7 reference genome (PF3D7_0323400). Non-synonymous SNPs were called at a minimum frequency of 0.02 (2%) and a minimum coverage of 1000 reads. As a control, PfRIPR was amplified from Pf3D7 genomic DNA and sequenced to confirm the absence of SNPs arising from PCR or sequencing errors. SNPs were individually inspected for final validation. Sequence analysis and SNP calling were independently performed by at least 3 individuals for each sample.

### Structural Modeling of SNPs

The structures of PfCyRPA-PfRIPR-Cy.003 (PDB ID: 8CDE) and PfRCR-Cy.003 (PDB ID: 8CDD) were downloaded from the Protein Data Bank (PDB). FASTA files were created of the reference PfRIPR amino acid sequence as well as PfRIPR sequence with individual SNPs (novel and previously identified). The amino acid sequence files were threaded onto the crystal structure of the PfRIPR-PfCyRPA-PfRH5 (PfRCR) complex and Pymol version 3.0.4 was used to plot the structural location of each SNP. The effect of SNPs on RIPR stability and RIPR-CyRPA binding energy were predicted by FoldX 4.0.

## Supplementary Materials

**Supplementary Table 1A:**
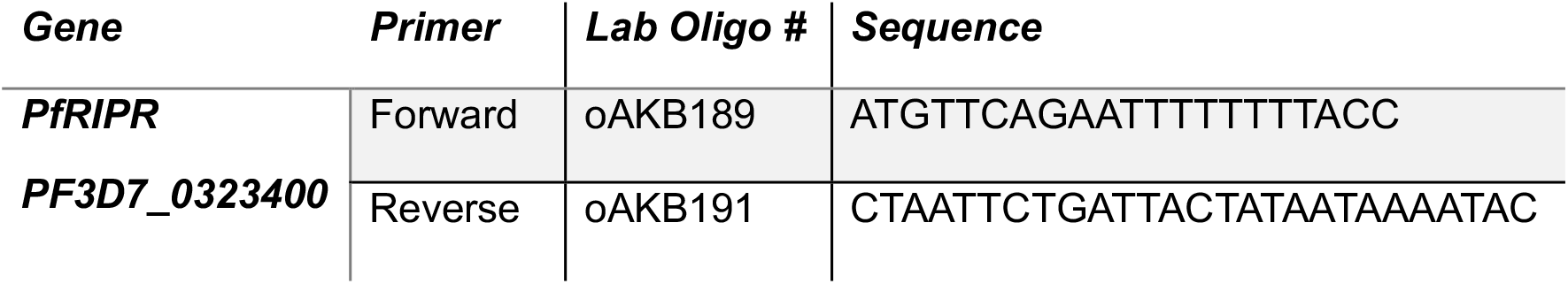
PfRIPR primers for full length sequence (2022 and 2023 samples)

**Supplementary Table 1B:**
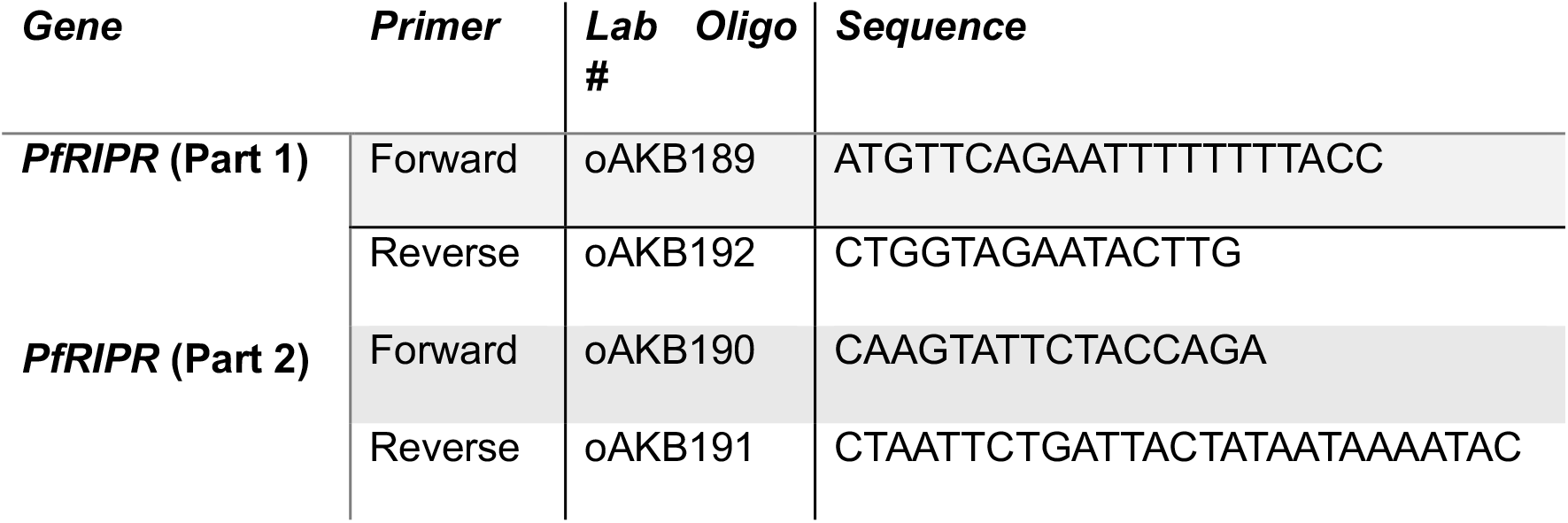
PfRIPR primers for partial sequence (2019 samples)

**Supplementary Table 2:**
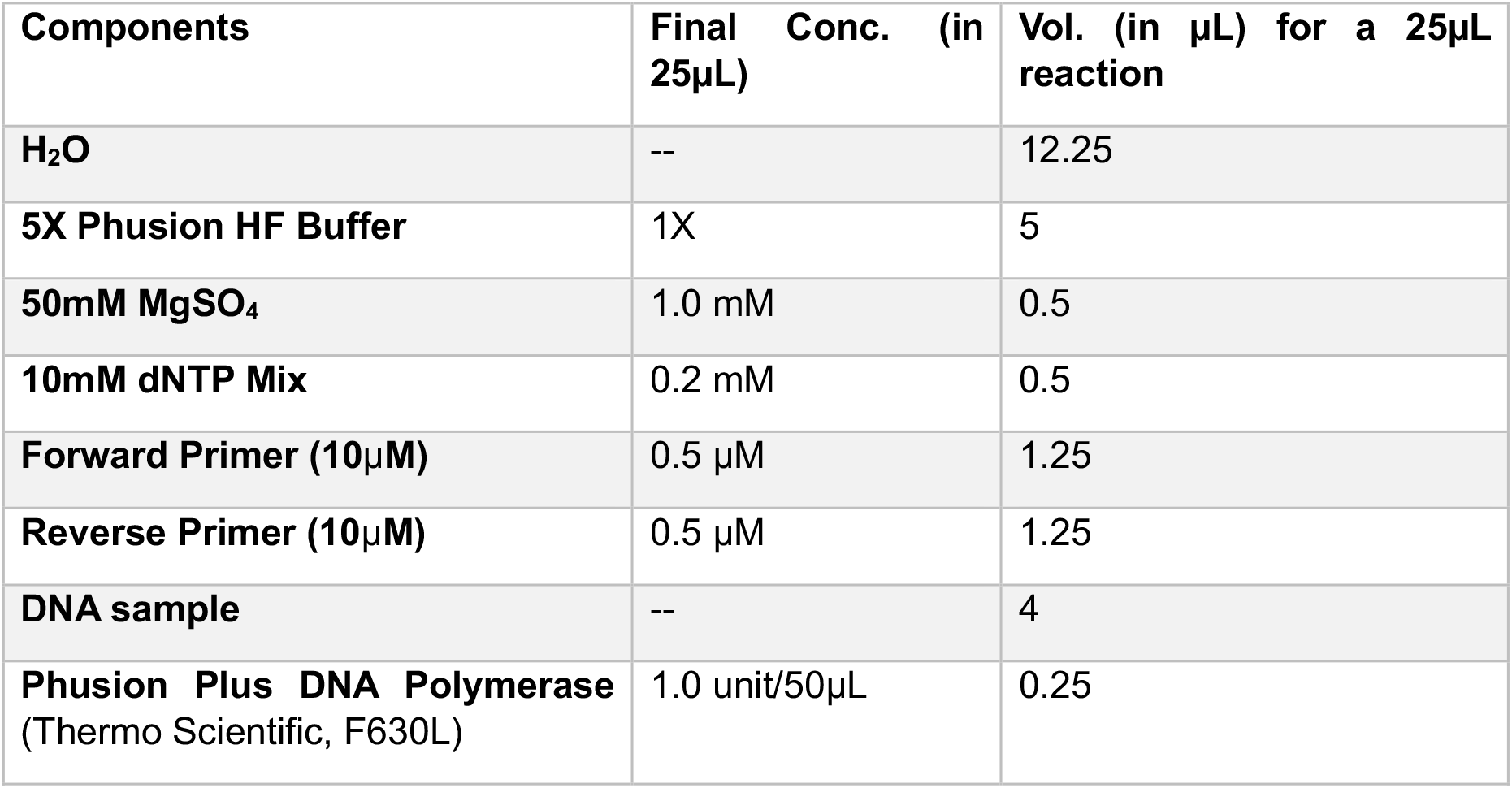
PCR Reaction Mix.

**Supplementary Table 3:** PCR program.

| Step | Cycle | Temperature (°C) | Time | Number of Cycles |
| --- | --- | --- | --- | --- |
| 1 | Initial Denaturation | 98 | 30 seconds | 1 |
| 2 | Denaturation | 98 | 10 seconds | 39 |
| 3 | Annealing | 57 | 45 seconds |  |
| 4 | Extension | 72 | 3 minutes |  |
| 5 | Final Extension | 72 | 5 minutes | 1 |
| 6 | Refrigeration | 12 | HOLD | --- |

**Supplementary Table 4:**List of all samples with identified SNPs, variant read frequencies, demographic and parasitological characteristics of the study participants.

Attached file: *Supplementary Table 4*.*xlsx*

**Supplemental Figure 1:**
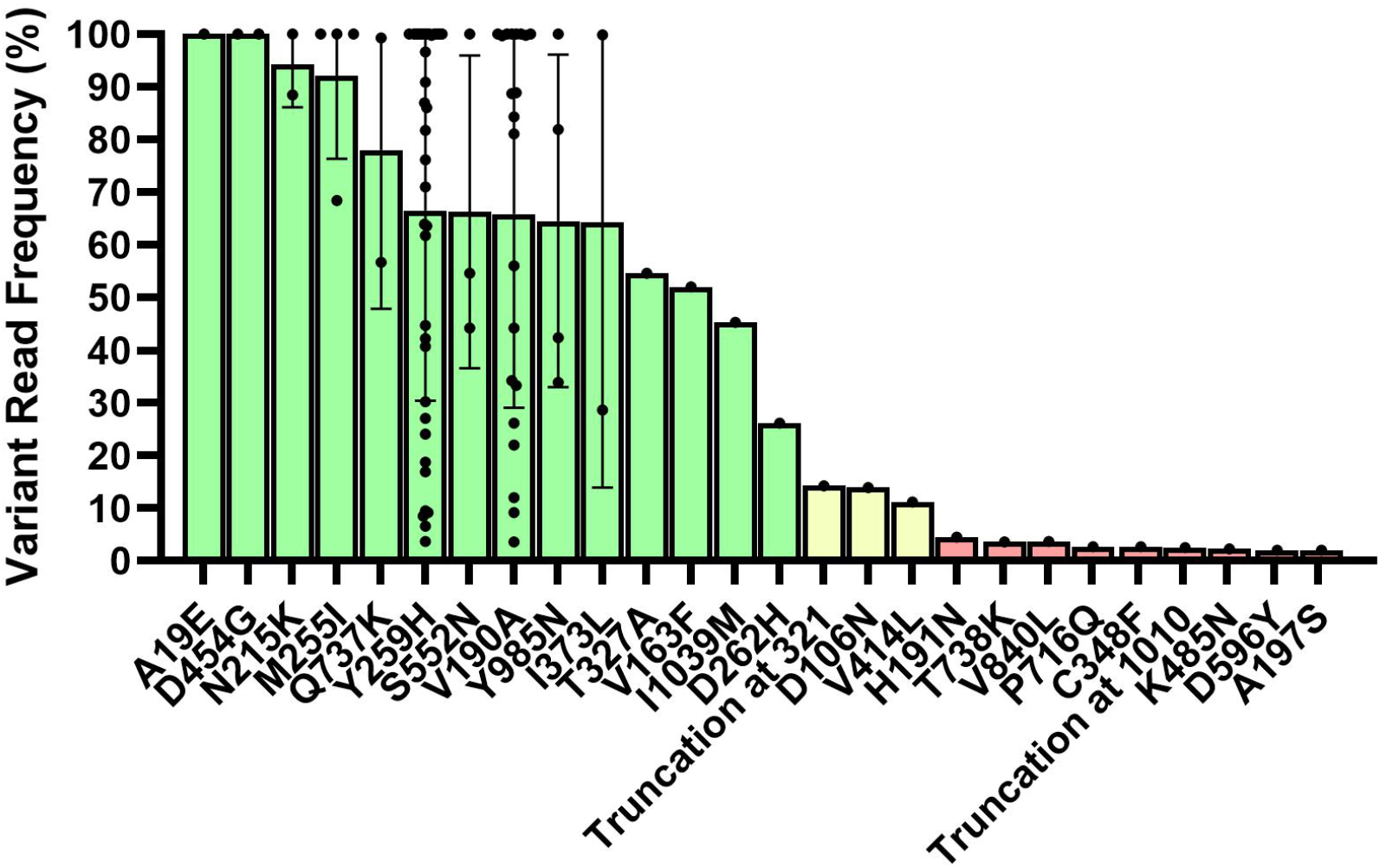
Variant read frequency of each nonsynonymous SNP identified. Each SNP is ranked by mean variant read frequency. Each dot represents the variant read frequency of a SNP in a single sample. Green bars represent high frequency (>25%), yellow bars represent intermediate frequency (5-25%), and red bars represent low frequency (2-5%).

